# Inpatient Skin-to-Skin Care Predicts 12-month Neurodevelopmental Outcomes in Very Preterm Infants

**DOI:** 10.1101/2023.04.06.23288260

**Authors:** Molly F. Lazarus, Virginia A. Marchman, Edith Brignoni-Pérez, Sarah Dubner, Heidi M. Feldman, Melissa Scala, Katherine E. Travis

## Abstract

**Objective:** Limited research links hospital-based experiences of skin-to-skin (STS) care to longer-term neurodevelopmental outcomes in preterm children. The present study examined relations between inpatient STS and neurodevelopmental scores measured at 12 months in a sample of very preterm (VPT) infants.

**Study Design and Methods:** From a retrospective study review of medical records of 181 VPT infants (<32 weeks gestational age (GA)) we derived the STS rate, i.e., the total minutes of STS each infant received/day of hospital stay. We used scores on the Capute Scales from routine follow-up care at 12 months as the measure of neurodevelopmental outcome (n=181).

**Results:** Families averaged approximately 17 minutes/day of STS care (2 days/week, 70 minutes/session), although there was substantial variability. Variation in STS rate was positively associated with outcomes at 12 months corrected age (*r* = 0.25, *p <* .001). STS rate significantly predicted 6.2% unique variance in 12-month neurodevelopmental outcomes, after controlling for GA, socioeconomic status (SES), health acuity, and visitation frequency. A 20-minute increase in STS per day was associated with a 10-point increase (.67 SDs) in neurodevelopmental outcomes at 12 months. SES, GA, and infant health acuity did not moderate these relations.

**Conclusion:** VPT infants who experienced more STS during hospitalization demonstrated higher scores on 12-month assessments of neurodevelopment. Results provide evidence that STS care may confer extended neuroprotection on VPT infants through the first year of life.

## Introduction

Caregiver-infant skin-to-skin (STS) care is a developmental care practice associated with numerous benefits to short-term health outcomes for infants born preterm.^1–4^ STS care positively impacts infant health outcomes (e.g. cardiorespiratory stability, growth, infection rates) and parenting practices (e.g. attachment, breastfeeding), which are, in turn, important predictors of neurodevelopment.^4–9^ However, direct evidence linking hospital-based experiences of STS to *longer-term neurodevelopmental outcomes* in children born preterm is currently lacking.^10^ Moreover, it is unknown whether STS care uniquely predicts outcomes over and above other clinical and socio-demographic factors, such as gestational age (GA), socioeconomic status (SES), neonatal health, or frequency of family visitation. Such data are needed to understand the utility of STS care for mitigating adverse neurodevelopmental sequelae associated with preterm birth.

We investigated the relation between amount of family STS care in the Neonatal Intensive Care Unit (NICU) and child neurodevelopmental outcomes at 12-months age (adjusted for degree of prematurity) in infants born very preterm (VPT). We hypothesized that amount of STS would significantly predict scores on a measure of child neurodevelopment at 12 months of age. We further explored this relationship after accounting for other predictors of neurodevelopmental outcomes, such as GA at birth, SES of the family, and the infant’s medical risk for adverse outcomes. We controlled for family visitation to assess the degree to which effects were specific to STS rather than reflective of general features of family involvement.^11^ We additionally controlled for prior neurodevelopmental scores at 6-months to assess the persistence of effects over time and to adjust for stable attributes of the child and family. Results shed light on the degree to which STS care may be neuroprotective and provide needed empirical justification for promoting institutional and social supports for STS care for infants born very preterm.

## Method

### Design

Data were collected during routine clinical care and retrospectively derived from the Electronic Medical Record (EMR). Participants were not required to give consent because this study was considered a minimal-risk retrospective chart review. Stanford University Institutional Review Board approved the experimental protocol (#IRB-44480).

### Sample

Infants met inclusion criteria if they were born very preterm birth (< 32 weeks) between 5/1/2018 to 6/15/2022, cared for at Lucile Packard Children’s Hospital, and returned to our clinic for follow-up at 12 months corrected age (*n=*181; 47% Female). Infants were excluded if they transferred into our NICU after the first 7 days of life (*n*=15), had a diagnosis of a genetic or congenital anomaly known to affect neurodevelopment (*n*=13), or were missing our outcome variable of interest (*n*=23). A protocol standardizing the execution and charting of developmental care, including STS, was fully operational by 5/1/2018. Therefore, only infants born after this date were included to ensure reliable and standardized tracking of STS care for the duration of hospitalization.

### Measures

#### Clinical and demographic measures

Demographic and clinical characteristics of infants were extracted from the EMR, including sex assigned at birth (male or female), GA (weeks), age at hospital admission (days), weight at birth (g), and length of hospital stay (days). We used insurance status (private versus public) to index family SES. In California, qualification for public insurance is calculated based on a family’s income-to-needs ratio and is thus, a reflection of family economic resources.

Information regarding four major comorbidities of prematurity was also extracted. Bronchopulmonary dysplasia (BPD) was defined as treatment with supplemental oxygen at 36 weeks postmenstrual age (PMA). Intraventricular Hemorrhage (IVH) was defined as the presence of a grade I or higher using the Papile classification system.^12^ Sepsis was defined as a positive blood culture or >7 days of antibiotics. Necrotizing enterocolitis (NEC) was defined as a diagnosis of medical or surgical NEC. A binary health acuity score was calculated to categorize infants into those with none versus one or more of these conditions.

#### In-hospital skin-to-skin care and family visitation

As part of routine daily charting at our hospital, all bedside nurses document in the EMR instances of family engagement in developmental care activities as they occur. Charting of developmental care activities notes the type (Skin-to-skin Care, Swaddled Holding, Touch, Massage, Music, Talking, and Singing), the approximate duration in minutes, and who is involved (mother, father, other family member, nurse, other staff member, or any combination of these). Starting May 2018, a developmental care protocol, the iRainbow^13^, was implemented and created standardized clinical criteria for STS delivery from staff and family members. Given our focus on STS, we specifically extracted STS instances and computed STS rate as the number of minutes of STS that each infant received/number of days of hospital stay (to account for individual variation in length of hospital stay). We computed two additional metrics of STS to further understand patterns of STS occurring in our unit. STS frequency captured how often families engaged in STS sessions as the number of instances of STS/number of days of hospital stay. STS duration captured the average length of STS sessions as the number of minutes of each STS session/number of instances of STS across hospital stay.

Bedside nurses also document any time there is a person at the bedside who is not clinical staff and note who is visiting (mother, father, other family member, or any combination of these). We defined family visitation as the total number of visitation instances by any family member/number of days of hospital stay (to account for individual variation in length of hospital stay).

#### Neurodevelopmental outcomes

Infants born < 32 weeks GA are eligible for developmental follow up as a part of California’s High-Risk Infant Follow Up program. Our metric of neurodevelopment was The Capute Scales Composite score^14^ assessed at 6- and 12-months age (adjusted for prematurity) and recorded in the EMR. The Capute Scales is a 100-item measure including a cognitive and language component. The Cognitive Adaptive Test (CAT) uses clinician observation and parental report to assess visual-motor problem-solving ability in standardized tasks (e.g., “pulls down a ring,” “releases one cube in a cup”). The Clinical Linguistic and Auditory Milestone Scale (CLAMS) uses parent report and (when possible) clinician observation to assess expressive and receptive language skills (e.g., “makes razzing sounds,” “orients to bell laterally”).

Quantitative developmental quotients (DQ = developmental age/adjusted age x 100) are derived separately for the CAT and the CLAMS and DQs are averaged to derive the composite score.

Age, adjusted for the number of weeks preterm at birth, was used to calculate developmental quotient scores. If infants were missing scores on either test a single score was used. A DQ ≤ 70 suggests delays. Prior work has demonstrated the Capute’s concurrent and predictive validity to other standardized assessments such as the Bayley Scales of Infant Development.^15^

#### Analytic strategy

All analyses were conducted in R version 4.2.2. We examined descriptive statistics for visitation frequency, each metric of STS, and neurodevelopmental scores. All variables were inspected for outliers tested for normality. Any variable with a value above or below three standard deviations from the mean was winsorized. Shapiro-Wilk tests identified STS rate and frequency as non-normal, so they were transformed using a base 10 log. Zero-order correlations inspected relations between predictors and 12-month neurodevelopmental scores. A hierarchical linear regression examined the relation between STS and 12-month neurodevelopmental scores when controlling for covariates. We chose GA, SES, and health acuity as covariates because we expected *a priori* that these factors would be associated with outcomes. We included visitation frequency as a covariate to distinguish the effects of STS from a general proxy of family presence or involvement. Additional analysis included 6-month scores as a covariate because earlier scores are likely to be related to later scores due to continuity across development and stable features of the child/family, thus controlling for 6 month scores allowed us to determine if there are extended benefits of STS care to child neurodevelopmental outcome a 12 months.

Follow-up analyses also examined STS frequency and duration as predictors of neurodevelopmental scores. Given that rates of developmental care are known to have been impacted by the pandemic,^16^ and the possibility that SARS-CoV2 pandemic (COVID) impacted follow up, we re-ran regression analyses including COVID as a covariate. COVID was defined as those whose entire hospital stay and follow up occurred prior to 3/8/2020 (89 infants; 44%), those who experienced their hospital stay prior to 3/8/2020 but were followed up on or after 3/8/2020 (9 infants: 4%), those who experienced some portion of their hospital stay on or after 3/8/2020 (106 infants; 52%). We also tested GA, health acuity, SES and birth during COVID as moderators of the relations between STS and 12-month outcomes. All significance levels were set at *p* < .05.

## Results

Table 1 summarizes clinical and demographic characteristics of the sample. Our sample included an equal number of females as males; half of the sample received public (rather than private) insurance. All children were born preterm with a mean GA of approximately 28 weeks and mean birth weight of approximately 1116 g. The mean length of hospital stay was about 2.6 months.

**Table 1.** Descriptive Statistics of the Sample (n=181)

|  | **M (SD) or n (%)** | **Min** | **Max** |
| --- | --- | --- | --- |
| Female (%) | 85 (46.96) | - | - |
| Public Insurance (%)<sup>1</sup> | 83 (45.86) | - | - |
| GA at Birth (weeks) | 28.26 (2.53) | 22.10 | 31.80 |
| Weight at Birth (g) | 1116.67 (389.70) | 410.00 | 2059.90 |
| Length of Stay (days) | 79.65 (43.88) | 23.30 | 240.21 |
| Health Acuity (%)<sup>2</sup> | 90 (49.72) | - | - |
| BPD (%) | 49 (27.07) | - | - |
| IVH (%) | 47 (25.97) | - | - |
| NEC (%) | 35 (19.33) | - | - |
| Sepsis (%) | 19 (10.50) | - | - |
| Visitation Frequency<sup>3</sup> | 0.99 (.45) | 0.06 | 2.54 |
| STS Rate<sup>4</sup> | 17.65 (16.62) | 0.00 | 83.38 |
| 6-Month Outcome<sup>5</sup> | 94.68 (14.00) | 39.42 | 135.00 |
| 12-Month Outcome<sup>6</sup> | 91.64 (14.93) | 52.00 | 136.00 |

<sup>1</sup> SES as defined by the percentage of families with public vs private health insurance.
<sup>2</sup> Health acuity score reflecting diagnoses of one or more of the following conditions: bronchopulmonary dysplasia (BPD), intraventricular hemorrhage (IVH), necrotizing enterocolitis (NEC), sepsis.
<sup>3</sup> Number of instances of visitation/number of days of inpatient hospital stay.
<sup>4</sup> Number of minutes of skin-to-skin care (STS)/number of days of inpatient hospital stay.
<sup>5</sup> 6-month developmental quotient scores on the Capute Scales (calculated using adjusted age) (n=166).
<sup>6</sup> 12-month developmental quotient scores on the Capute Scales (calculated using adjusted age).

Table 1 also reports visitation frequency and STS rates. A visitation frequency of 1.0 would indicate that visitation occurred daily. Here, families visited almost every day, on average, though some families visited more than twice a day. Over their entire stay, on average, infants received 17 minutes/day of STS. A total of 14 infants (8%) received more than 50 minutes/day; 13 families (7%) did not engage in any STS. Families engaged in STS less than 2 days/week (M = 0.25, SD = 0.23) on average. Instances of STS lasted a mean of 70 minutes (SD = 15.78), with a range of 30 to 123 minutes/session. Infants from higher SES families experienced more minutes of STS per day (M = 23.18, SD = 17.57), compared to those from low SES families (M = 11.12, SD = 12.71) (*t* = 5.34 *p* < .001). Visitation frequency was positively associated with STS rate (*r* = 0.38, *p* < .001); families who visited more frequently tended to perform STS for more minutes/day.

Due to variability in available clinical developmental assessments, 23 children returned for a 12 month visit at our High Risk Follow Up Clinic but were not assessed using the Capute. T-tests and chi-square tests indicated no significant health or sociodemographic differences between infants who were assessed using the Capute (n=181) and those who came to the clinic but did not have a reported Capute score (n=23) at the 12-month visit. Of the 181 infants with 12-month scores, 92% (n=166) had available 6-month Capute scores and were thus included in analysis controlling for 6-month outcomes. There were no significant clinical or demographic differences between the infants who had both 6 and 12-month scores (n=166) and those who did not have Capute scores at 6 months (n=15). Of those who were tested using the Capute, no infants had missing scores on the CLAMS but 18% (n=33) of infants were missing CAT scores at 12 months and 12% (n=20) were missing CAT scores at 6 months. Infants who had missing scores on the CAT were not demographically or clinically different than those with complete scores at either time point although they were more likely to have undergone assessment in the acute phase of the SARS-CoV2 pandemic (χ² = 20.627, p < 0.001). Critically, there were also no statistical differences in Capute scores between those whose scores were calculated using both CAT and CLAMs compared to those using only CLAMS scores.

The average adjusted age at the initial follow-up visit was 5.50 months (SD = 0.82) and 13.32 months (SD = 1.46) at the second visit. As shown in Table 1, developmental quotients varied substantially across in the infants. At 6 months, 4 (3%) children had scores suggesting developmental delay (< 70); at 12 months, 10 (6%) children fell into this range. STS was associated with 6-month scores such that infants who experienced more minutes of STS during their hospital stay tended to have higher scores at 6 months of age (*r* = 0.19, *p* = .02). STS was related to 12-month scores such that infants who experienced more minutes of STS during their NICU stay had higher neurodevelopmental scores (*r* = 0.25, *p* < .001).

Table 2 documents the unique contribution of STS rate to neurodevelopmental outcomes. Model 1 shows that the covariates accounted for approximately 7% of the variance in child outcomes. Model 2 demonstrates that STS uniquely predicted 12-month neurodevelopmental scores after GA, SES, health acuity, and family visitation, with STS accounting for approximately 6% unique variance. A 1% increase in STS was associated with .09-point increase in 12-month scores, thus on average, a 20-minute increase in the amount of average daily STS was associated with a 10.09-point increase in scores on 12-month neurodevelopmental assessments (B=8.90, 95% CI 3.94–13.87), more than 2/3rds of a SD increase. Model 4 demonstrates that STS was also uniquely predictive of 12-month scores even when including 6- month scores as a covariate (B=7.03, 95% CI 1.99–12.07). Figure 1 illustrates the association of STS with neurodevelopmental scores, after adjusting for covariates.

**Figure 1.**
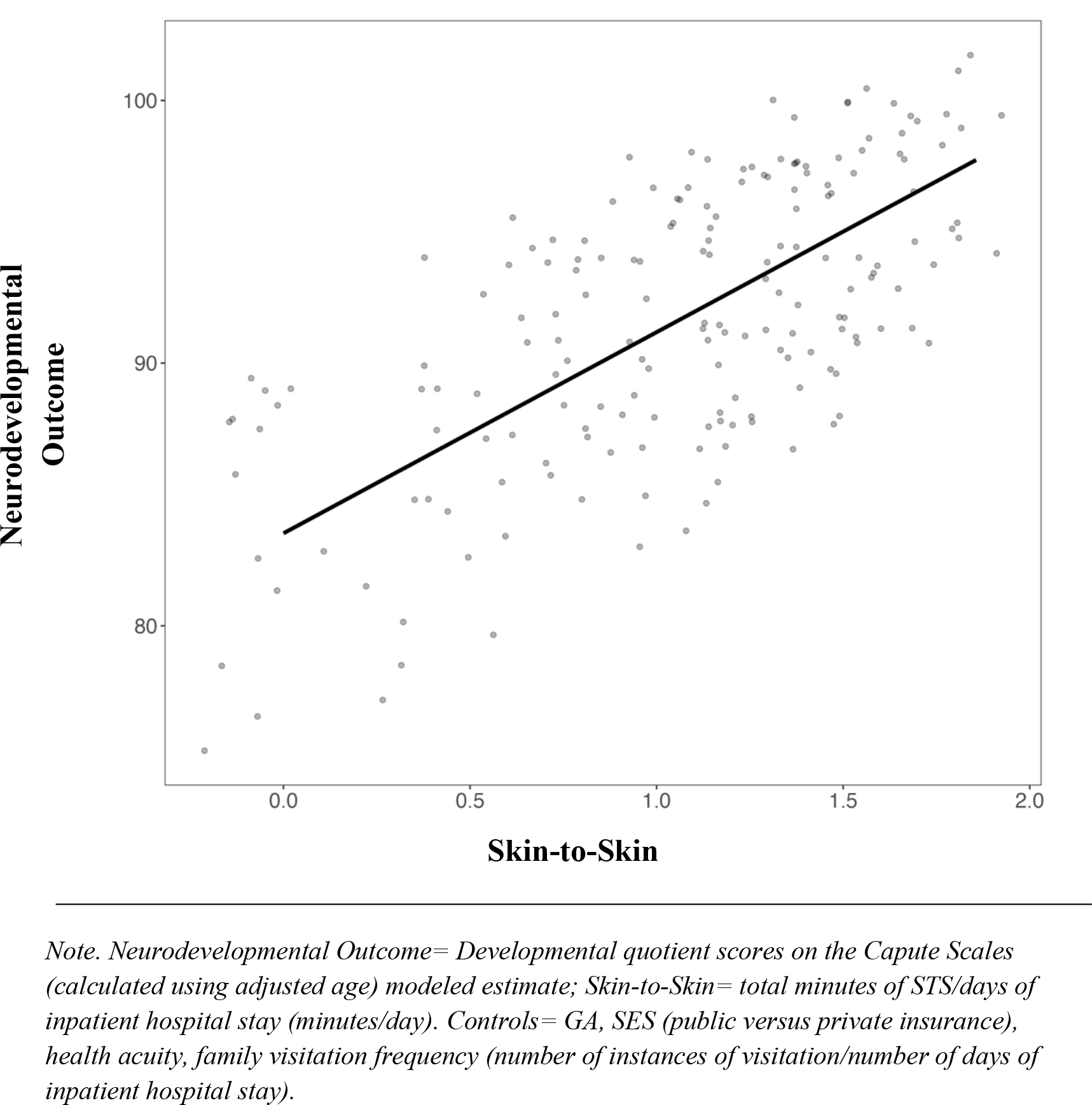
Modeled estimate of the association between skin-to-skin and neurodevelopmental outcomes at 12 months, after covariates.

**Table 2.** Multiple regression models (unstandardized coefficients) predicting 12-month neurodevelopmental scores.

|  | Model 1 | Model 2 | Model 3 | Model 4 |
| --- | --- | --- | --- | --- |
|  | B (95% CI) | B (95% CI) | B (95% CI) | B (95% CI) |
| Gestational Age | 0.55<br>(-0.47 – 1.57) | 0.57<br>(-0.43 – 1.56) | 0.37<br>(-0.66–1.40) | 0.39<br>(-0.62 – 1.40) |
| SES<sup>1</sup> | -2.01<br>(-6.30 – 2.27) | 1.23<br>(-3.30 – 5.76) | -2.47<br>(-6.70–1.77) | 0.15<br>(-4.41 – 4.70) |
| Health Acuity<sup>2</sup> | -5.72*<br>(-10.95 – -0.49) | -4.63<br>(-9.74 – 0.48) | -6.28*<br>(-11.45–1.11) | -5.63*<br>(-10.3 -0.6) |
| Visitation<sup>3</sup> | -0.24<br>(-5.09 – 4.60) | -3.45<br>(-8.47 – 1.57) | 1.31<br>(-3.51 – 6.12) | -1.16<br>(-6.19 – 3.88) |
| 6-month Score<sup>4</sup> | - | - | 0.33***<br>(0.18–0.49) | 0.29***<br>(0.14 – 0.45) |
| STS Rate<sup>5</sup> | - | 8.90***<br>(3.94 –13.87) | - | 7.03**<br>(1.99 – 12.07) |
| Observations | 181 | 181 | 166 | 166 |
| R<sup>2</sup> | 0.07* | 0.13*** | 0.19*** | 0.23*** |
| r<sup>2</sup> Δ<sup>6</sup> | - | 0.06*** | - | .04** |

<sup>1</sup> Private versus public health insurance.
<sup>2</sup> Diagnosis of one or more of the following conditions: bronchopulmonary dysplasia (BPD, intraventricular hemorrhage (IVH), necrotizing enterocolitis (NEC), sepsis.
<sup>3</sup> Number of instances of visitation/number of days of inpatient hospital stay.
<sup>4</sup> Developmental quotient scores on the Capute Scales Cognitive Adaptive Test, adjusted for prematurity.
<sup>5</sup> Number of minutes of Skin-to-skin (STS)/number of days of inpatient hospital stay.
<sup>6</sup> R<sup>2</sup> Δ for Model 2 is in reference to Model 1 R<sup>2</sup>; R<sup>2</sup> Δ for Model 4 is in reference to Model 3 R<sup>2</sup>.
\* $p < .05$ ; \*\* $p < .01$ ; \*\*\* $p < .001$ .

Birth during COVID as a covariate did not alter the amount of variance in neurodevelopmental scores predicted by STS rate (B=9.20, 95% CI 4.23–14.18, STS R^2^ Δ = 0.06). In addition, GA, SES, health acuity and birth during COVID did not moderate these patterns (all *p* > .1), suggesting that relations between STS and 12-month neurodevelopmental outcomes were similar across clinical, SES, and birth groups.

Additional analysis demonstrated that STS frequency and duration similarly predicted 12- month cognitive outcomes, controlling for GA, SES, health acuity, and family visitation. STS frequency accounted for 4% unique variance. A 20 percent increase in the frequency with which families engaged in STS corresponded to a mean 9.6-point increase in neurodevelopmental scores at 12 months (B=48.43, 95% CI 13.87–82.99). STS duration accounted for 3% unique variance. An increase in the duration of STS by 20 minutes/session was associated with a 3.74- point increase, in neurodevelopmental scores (B =.19, 95% CI 0.03–0.34), more than a of a quarter of a SD increase.

## Discussion

This study contributes to a growing body of evidence that STS care may serve as a neuroprotective strategy for preterm infants at risk of developmental delay. Consistent with our hypotheses, infants who experienced more STS over their hospital stay had higher scores on a standardized measure of neurodevelopmental abilities at 12 months. The magnitude of our observed effect size (two thirds of a standard deviation in neurodevelopmental scores) is noteworthy given that meta-analysis of other interventions specifically targeted at cognition have shown an average of .42 standard deviations in score improvement.^17^ These relations remained when controlling for clinical, demographic, and developmental predictors that may influence neurodevelopmental abilities. Additionally, relations between STS and neurodevelopmental scores did not differ based on infants’ GA at birth, SES, or health status, suggesting that STS may be beneficial for children from a range of demographic and health backgrounds. STS predicted outcomes over and above frequency of family visitation, suggesting that specifically having families actively engage in STS activities may contribute to positive neurodevelopmental outcomes to a greater extent than simply encouraging families to visit the hospital without engaging in STS. Finally, we found that STS care contributed to child outcomes even after controlling for prior neurodevelopmental scores, suggesting that the effects of STS care during hospitalization are long lasting and persist at least until 12 months beyond factors that are attributable to stable features of the child and/or family context.

The present findings are generally consistent with previous observational studies showing positive relationships between amounts of parental STS and gross motor development,^11^ as well as amounts of parent holding activities and emergent language abilities in preterm infants.^18^ Additionally, work from Lester and colleagues^19^ demonstrated an association between maternal engagement in the NICU and cognitive scores at 18 months which was driven by STS care. The current study adds to this work by emphasizing the specificity of in-hospital experience of STS for predicting later neurocognitive outcomes in preterm infants. The design and conception of the present study also closely resembles that of Gonya and colleagues,^20^ who reported positive relations between parental STS and cognitive outcomes. However, the associations did not reach statistical significance in their study, possibly because their measure of STS was tracked at the hour level and developmental outcomes were categorized into high/low binary groups. A strength of the current study is the specificity and robustness of our minute-level STS metrics, and continuous measurement of cognitive scores, distinctions that give us more power to detect significant associations.

Prior evidence for the long-term protective properties of STS on neurodevelopment comes from randomized control trials.^21,22^ Such studies addressed the issue of causality and established efficacy of STS as a medical intervention for supporting longer-term outcomes. However, such studies do not necessarily reflect the real-life variation of STS that exists outside of stringent experimental settings. Using a retrospective cohort design, our study was able to examine variations in patterns of family STS that occurred naturally during hospitalization, independent of any explicit study involvement. Therefore, our results complement these earlier efficacy studies by documenting the effectiveness of STS as a potential clinical intervention for supporting neurocognitive development. Our findings add to a growing body of literature documenting the neuroprotective benefits of family-centered care practices for supporting health outcomes of preterm infants.^23,24^

The level of explanatory power captured in our study is particularly noteworthy, given the relatively small amounts of STS families provided. On average, infants were exposed to STS for an average of less than 20 minutes/day, 2 days per week, and for sessions that lasted about one hour. Given that transitioning babies from their cribs may be potentially stressful and disruptive, and considering infant’s natural sleep cycles, our current hospital protocols recommend that STS sessions should last *at least* 60 minutes. Many sessions failed to reach this 1-hour benchmark. It is impressive that variation in levels of STS activity even in these low ranges can nevertheless contribute predictive utility to neurodevelopmental outcomes at 12 months of age. Future research should explore the possibilities of a threshold-dose effect or if amounts of STS at higher baseline levels show similar or possibly even greater effects.

Several mechanisms may be at play to explain the observed relations between STS and better neurodevelopmental scores. First, STS has been related to stress reduction and autonomic regulation^25,26^ both of which affect neurodevelopment.^27,28^ Additionally, STS may provide a formative bonding experience for infants and their caregivers,^6^ which in turn, may facilitate dyads’ capacities for exploration, teaching, and shared attention later in infancy.^29^ STS may also directly contribute to brain maturation and emergent neurocognitive abilities by providing non-noxious sensorimotor neural stimulation.^30^ Future studies should investigate these potential mechanisms more thoroughly.

This study had limitations. First, because this was a retrospective study, we were limited to data collected as part of routine clinical care. We did not have information about other factors that may have contributed to infants’ neurodevelopmental outcomes, such as caregiver education level or caregiving activities in the home. We cannot rule out the possibility that caregivers who engaged in more STS may have also engaged in more stimulating activities in the home environment after hospital discharge. However, in this scenario, caregiver involvement in STS may serve as an important early clinical marker for parental engagement helpful in identifying infants or families who may require assistance or interventions that support neurodevelopment in the home environment. We also did not have information about factors that may have impacted caregiver’s ability (e.g., access to paid family leave, childcare for other children, transportation) to perform STS. More research is needed to understand whether modifying such factors can be beneficial in promoting access to STS. An additional limitation was that our measures of STS relied on clinical charting. However, clinical staff members are specifically trained and have been routinely charting developmental care practices since before the onset of this study. We did not rely on parental report as in previous studies^18,31,32^ which can be prone to reporting biases.

The retrospective cohort design conferred a significant strength: generalizability. Because active participation was not required, our sample captured occurrences of STS as implemented *in situ* in the daily lives of all families in the NICU, including many individuals who often go unstudied due to self-selection bias or financial/social circumstances.

## Conclusion

Overall, our findings argue in favor of increasing institutional and societal supports to promote opportunities for families to engage more directly and regularly in the care of their preterm infants. Many parents of hospitalized infants feel that their role as a parent is diminished because they lack the agency and expertise to care for their child.^37^ Increased institutional support and education about the benefits of family-administered STS may help families in the NICU feel more empowered in their unique ability to help their babies health both in the short- and long-term. Clinical staff may be reluctant to support family engagement in STS because of the misconception that the benefits of STS have not been based on scientific evidence.^38^ The present study may help to dispel this notion. Increased societal support, such as improved paid parental leave policies, would also be an important component of providing families with more equitable opportunities to provide STS to their hospitalized infants. Such supports are likely to be important factors in mitigating the negative long-term consequences of preterm birth on neurodevelopment that disproportionately affect lower-income and non-white families.^39,40^ The results of this study contribute to growing evidence for the broad implementation of in-hospital STS as a low-cost, family-centered intervention to promote short- and long-term benefit in very preterm infants.

## Data Availability

Deidentified individual participant data will be made available upon request.

## Data Sharing Statement

Deidentified individual participant data will be made available upon request.

## Abbreviations

Skin-to-skin (STS); Very Preterm (VPT); Electronic Medical Record (EMR); Socioeconomic Status (SES); Gestational age (GA); Standard Deviation (SD); Lucile Packard Children's Hospital (LPCH); Neonatal Intensive Care Unit (NICU); Cognitive Adaptive Test (CAT); Clinical Linguistic and Auditory Milestone Scale (CLAMS).

